# Epidemiology of Extended-spectrum beta-lactamase-producing *Escherichia coli* at the human-animal-environment interface in Wakiso district, Uganda

**DOI:** 10.1101/2022.11.12.22282228

**Authors:** James Muleme, David Musoke, Bonny E. Balugaba, Stevens Kisaka, Frederick E. Makumbi, Esther Buregyeya, John B. Isunju, Wambi Rogers, Richard K. Mugambe, Clovice Kankya, Musso Munyeme, John C. Ssempebwa

**Affiliations:** Department of Disease Control and Environmental Health, Makerere University School of Public Health, Uganda; Department of Biosecurity Ecosystems and Veterinary Public Health, Makerere University College of Veterinary Medicine Animal Resources and Biosecurity, Uganda; Department of Disease Control, University of Zambia, Zambia; Department of Epidemiology and Biostatistics, Makerere University School of Public Health, Uganda; Clinical Laboratories, Mulago National Referral Hospital, Uganda

**Keywords:** Extended spectrum beta lactamase, Escherichia coli, farming, one health, interface, transmission

## Abstract

**Background:** Extended-spectrum beta-lactamase-producing *Escherichia coli* (ESBL-PE) represents a significant global public health concern. Much as humans, animals and environments harbor ESBL-PE, its epidemiology in Uganda is still not well understood. This study explains the epidemiology of ESBL-PE using the one health approach in selected farming households in Wakiso district, central Uganda.

**Methodology:** Environmental, human, and animal samples were collected from 104 households. Additional data were obtained using observation checklists and through interviews with household members using a semi-structured questionnaire. Surface swabs, soil, water, human and animal fecal samples were introduced onto ESBL chromogenic agar. The isolates were identified using biochemical tests and double-disk synergy tests. To assess associations, prevalence ratios (PRs) were computed using a generalized linear model (GLM) analysis with modified Poisson and a log link with robust standard errors in R software.

**Results:** A total of 82.7% (86/104) households had at least one positive ESBL-PE isolate. The overall prevalence of ESBL-PE at the human-animal-environment interface was approximately 25.0% (95% CI: 22.7-28.3). Specifically, animals, environment and humans had an ESBL-PE prevalence of 35.4%, 5.8%, and 45.4% respectively. Having visitors (adj PR= 1.19, 95% CI: 1.04-1.36), utilizing veterinary services (adj PR= 1.39, 95% CI: 1.20-1.61) and using animal waste for gardening (adj PR= 1.29, 95% CI: 1.05-1.60) were positively associated with ESBL-PE contamination. However, covering the drinking water container with a lid (adj PR= 0.84 95% CI: 0.73-0.96) was associated with absence of ESBL-PE.

**Conclusion:** There is wider dissemination of ESBL-PE in the environment, humans, and animals, indicating poor infection prevention and control (IPC) measures in the area. Improved collaborative one health mitigation strategies such as safe water chain, farm biosecurity, household and facility-based IPC measures are recommended to reduce the burden of antimicrobial resistance at community level.

## Background

If no appropriate measures are taken, it is projected that approximately 10,000,000 deaths and about US$100 trillion economic loss will occur per year by 2050 because of antimicrobial resistance [1].

Infections caused by Extended Beta Lactamase producing Escherichia coli (ESBL-PE) have been implicated in severe disease outbreaks globally [1,3,4]. Studies have reported that the resistance rates in E. coli ranged from 0–87% (to third generation cephalosporins) to 0–98% (to fluoroquinolones) [5,6]. In Tanzania, deaths due to sepsis among neonates have been attributed to ESBL infections [7]. A review by Martischang and colleagues in 2020, reported that the prevalence of ESBL-PE co-carriage ranged from 8% to 37% at the household level [8]. This implies that such infections are emerging “One Health” threats compromising the safety and health of humans, animals, as well as the purity of the environment globally. Even though irrational drug use has been pointed as an internationally recognized cause of intrinsic antimicrobial resistance (AMR), the relationships and associated dynamics among humans, animals and environment are increasingly being pinned as major phenomena for the registered global AMR burden [6,9] To understand the dynamics of the dispersal of ESBL-PE into natural environments beyond human and domestic animal population, it is important to keep in mind the general E. coli population as well. E. coli is ubiquitous, and asymptomatically colonizes the gut of birds and mammals.

The cause of human ESBL-PE colonization is still contentious since different studies started recognizing the role of the environment and animals in the development, spread and spillover of antimicrobial resistant pathogens (ARPs) [10,11,12]. On the other hand, studies have also reported that this relationship could be ambidirectional suggesting that humans could also spread the ESBL-PE to animals and the environment [8]. In addition, the inter-species transmission has also been reported globally (i.e., humans-humans and animals-animals) [9,13]. Indeed, there is a lot of undetected community carriage and transmission of ESBL-PE at the human-animal-environment interface [15] even though there is inadequate local evidence for this. Such insufficient evidence may not competently inform prevention and control strategies including policies to govern management of AMR in the LMICs [16,17].

Following the global call for management of AMR (WHO “tricycle protocol”), the government of Uganda developed the National Action Plan (NAP) for AMR to protect human, animal, and environmental health [18]. This plan also identifies a critical information gap regarding AMR especially at the Human-animal-environment interface. In areas like Wakiso district, an urban farming area with a high level of antimicrobial agent use, AMR is not uncommon [18, 19]. Coupled with having many livestock and humans, it presents a high opportunity for human-animal interaction hence providing a good platform to study ESBL-PE transmission dynamics. In response to the WHO “tricycle protocol” and the Uganda NAP, our study was formulated aiming at describing the epidemiology of ESBL-PE at the human-animal-environment interface in a local peri-urban farming community within Wakiso district in central Uganda.

## MATERIALS AND METHODS

### Study design and area

We conducted a cross-sectional study within farming communities of Wakiso district (00 24N, 32 29E). Wakiso district is the most populated district in Uganda with a household size of 4.7. It is also one of the districts with the highest number of livestock in Uganda [1]. The district has 503,442 households, 36% of these are engaged in both crop and livestock production [2]. The district is estimated to have these species of livestock: cattle (114,769), goats (132,964), sheep (27,542), pigs (199,962), chicken (2,783,509), ducks (33,350) and turkeys (4,852) [3]. Out of the total land area of 280,772.3 hectares, approximately 97,166.3 hectares (34.61%) are being used for agricultural production. Such demographic dynamics create a high chance of human-animal interaction.

### Sample size determination

Using Steve Bennett’s formula of cluster sample size calculation [4], 104 households were targeted as a primary unit of sampling. For data and sample collection, we adopted the modified Daniel’s formula [5], for categorical data and obtained a sample size of 840 samples (i.e., 280 for humans, 280 for animals and 280 for the environment). At each household, a four human samples, 4 animal samples and 4 environmental samples were picked for uniformity making a total of 1248 samples all together. After thorough quality checks, a total of 988 samples passed and these proceeded to the next level of laboratory analysis. Animal samples (fecal per rectal), human samples (urine and stool) and environmental samples (doorknobs, soil, animal feeding equipment and water) were picked from the same household.

### Study population and sampling procedure

Following a multi-stage sampling approach (Figure 1), our secondary sample (human, animal, and environment) was reached. All random sampling phases were conducted using a freely online available random number generator (www.random.org?). Lists of Sub counties, parishes were obtained from the district veterinary office. Briefly, Wakiso district was purposively selected due to its unique presentation (see section on “Study design and area”). A total of 8 Sub counties were purposively (livestock keeping) selected with the help of a veterinary officer and from these 50% were randomly selected. From each Sub County, 50% parishes were randomly selected while villages with animals, and have history of diarrhea episodes were purposively selected with the help of local chairpersons. The households involved in the study needed to possess more than four animal species and at least two members in the household. The household was our primary sampling unit whereas humans, animals and environment comprised of the secondary sampling unit. In addition, the household also served as a unit of analysis for the associations among humans, animals, and environment as far as the epidemiology of ESBL-PE is concerned.

**Figure 1:**
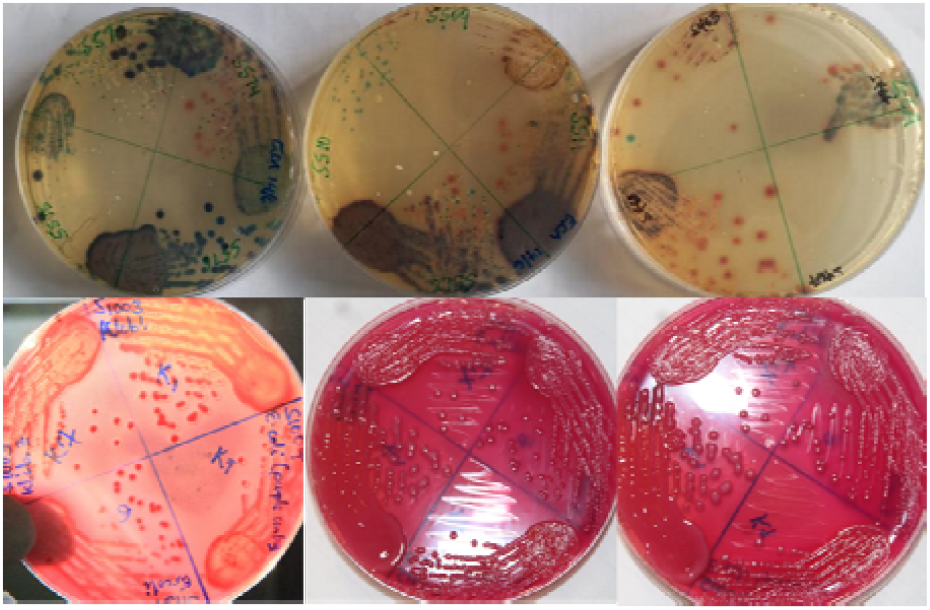
Primary cultures (upper row) and sub-cultures (lower row)

### Sample and data collection

A total of 104 farming households were recruited between March and July 2022. Briefly, household related data, human sample donor data, environmental inspection data, and animal characteristics data were collected at each household using a kobo collect based mobile application. A pre-tested semi-structured questionnaire was administered to household members who had experience with the key activities around the home.

One individual in a selected household was requested to provide a fecal and urine sample. A paper towel provided by the study teams and a sterile stool container with scoop was used by the participant. Mid-stream urine was also provided by the participants during sampling. Parents always supported their children to collect the samples after a thorough consent and assent process. Additionally, animal samples were picked per rectal from a minimum of 4 animals within the sampled household. Environmental samples including soil, water for domestic use, swabs from animal feeding equipment and doorknobs were also picked. The sample containers were labelled with the unique code that represented the household and the sample category. Within four hours of sample collection samples were delivered to the Microbiology Laboratory at the College of Veterinary Medicine Animal Resources and Biosecurity, Makerere University, in Ziploc bags under ice.

### Laboratory procedures

Sample processing was done by aseptically transferring sample inoculum on to freshly prepared ESBL chromogenic agar (Condalab 2062) which is used for detection of gram-negative bacteria producing extended spectrum Beta lactamase.

### Agar preparation and plating

Freshly incubated ESBL chromogenic agar containing ESBL supplement (condalab6042) having inhibition and selective agents was used [6]. Water and urine samples were first concentrated by centrifuging at 3000 rpm for 5 minutes prior to being introduced onto the agar. Soil samples and stool from Shoats were first emulsified/ suspended in 9mls of peptone water prior to plating. The plates were then incubated at 37□°C for 24 hours at ambient humidity and air conditions. Presumptive ESBL-PE appeared as pink, medium sized, raised and moist colonies (Figure 1).

### Identification and confirmation

*E. coli* isolates were further confirmed by biochemical tests following standard operating procedures [7]. *E. coli* was confirmed in case the isolate was positive for indole production, methyl red, motile, and negative for citrate utilization, urease production, and Voges-Proskauer. Those isolates with reduced susceptibility to cefotaxime (≤ 27□mm) and ceftazidime (≤ 22□mm) were confirmed for ESBLs-production using the modified double disk synergy (MDDS) method [19]. Briefly, after inoculation of the suspension onto Muller-Hinton agar (MHA), a disk of amoxicillin + clavulanic acid (20/10□μg) was placed in center of the plate and then the disks of cefotaxime (25□μg) and ceftazidime (30□μg) were placed at 20□mm from the central disk on the same plate. The plates were then incubated at 37□+C for 24□hrs and examined for an enhancement of inhibition zone of the β-lactam drugs caused by the synergy of the drugs and was interpreted as either being positive or negative for ESBLs-production (Figure 2).

**Figure 2:**
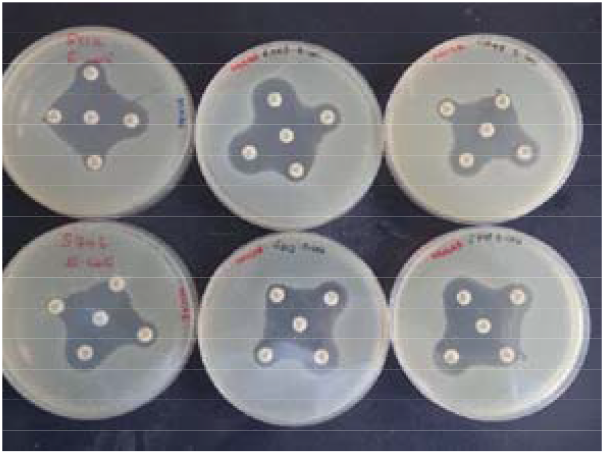
Modified Double Disc Diffusion test

### Study variables

Key independent variables for which data collected included individual demographics, household practices, animal husbandry practices, water, sanitation, and hygiene among other risk factors for occurrence of ESBL-PE in communities. The dependent variable was the ESBL-PE status (positive or negative) of the household as defined by the status of the respective humans, animals, and the environment.

### Data analysis and management

All data was cleaned for any missing variables or laboratory result to have a complete dataset. A new column was added for the outcome variable (laboratory outcome for each sample i.e., positive, or negative for ESBL-PE). This procedure was conducted for all data from the humans, animals, and those from the environment therefore this study had four datasets. All data analysis procedures were done in R version 4.2.1. Selected variables were picked from each individual dataset (human, animal, and environment) and merged to the main household dataset. Summary statistics were run, percentages and frequencies were reported in tables. Prevalence was defined as the total number of positive samples out of the total number of samples under consideration. Bivariate and multivariate robust modified Poisson regression were performed to generate prevalence ratios (PR) and respective 95% confidence intervals.

### Quality control and assurance

A trained research assistant collected all the samples and corresponding data. All samples were verified to confirm the labeling, type, adequate volume, and integrity at the laboratory. Media quality control was also performed by inoculating standard bacterial strain of *E. coli* ATCC 25922 which is ESBL negative and *Klebshiella Pneumoniae* ATCC 700603 which is ESBL positive. The media quality control form was scored PASS in case the *E. coli* failed to grow and *K. pneumoniae* grew.

### Ethical considerations

The Makerere University School of Public Health (SPH) institutional review board (IRB), the National Council for Science and Technology of Uganda (UNCST) and the district officials (head of administration – Chief administrative officer, District veterinary officer, District health officer) all ruled for the study and awarded us letters of support to that effect allowing us to conduct the study in Wakiso district, central Uganda. The study obtained ethical approval from Makerere University School of Public Health Institutional Review Board (SPH-2021-167) and the Uganda National Council for Science and Technology (HS1919ES). In addition, we obtained permission to conduct the study from Wakiso district headquarters (Chief administrative officer and district health and veterinary offices). Assent and consent forms were issued to the participants prior to their involvement into the study. Formal written consent was obtained from the parent/guardian before any child would be involved in the study. In addition, each of the abled child provided assent after being explained to the purpose of the study and its related processes. All ethical issues, and confidentiality were followed as guided by the Helsinki declaration.

## RESULTS

### Characteristics of the households involved in the study

This study was conducted among 104 households from four sub counties in Wakiso district. More than 60% (65/104) of the households had (5-9 members). Similarly, more than half of the sampled households had household heads who had attained primary school as the highest level of education, 71.1% (74/104) of the total households were male-headed. In addition, 80% (83/104) of the households had a shared water source between humans and animals. The detailed household characteristics are shown in Table 1.

**Table 1:**
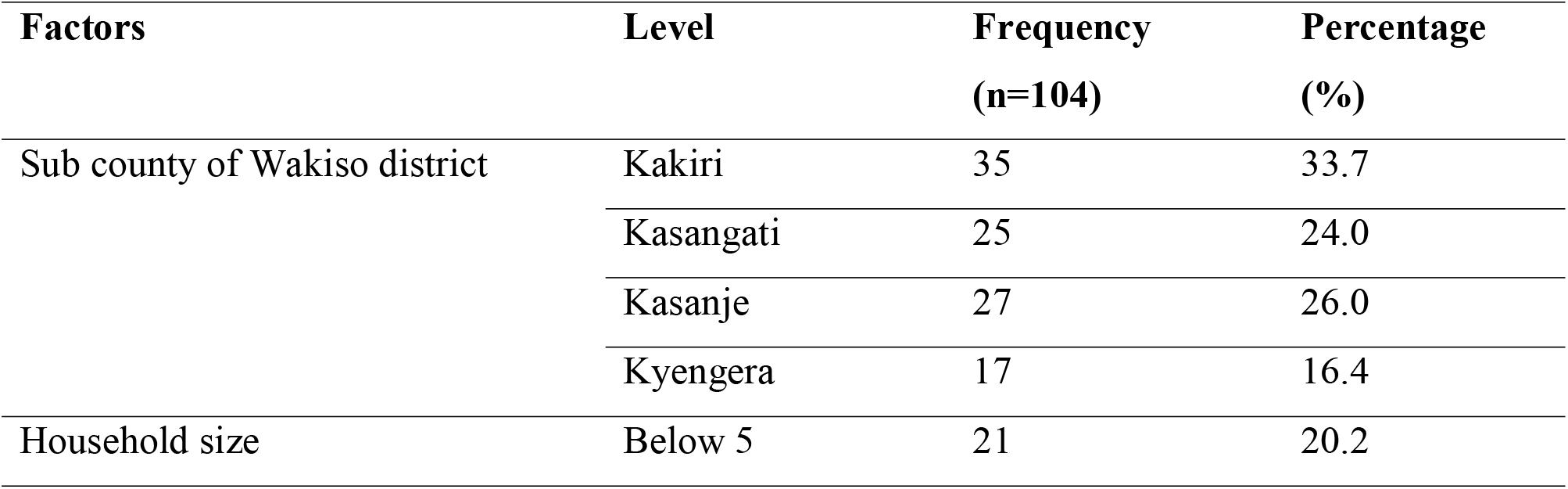
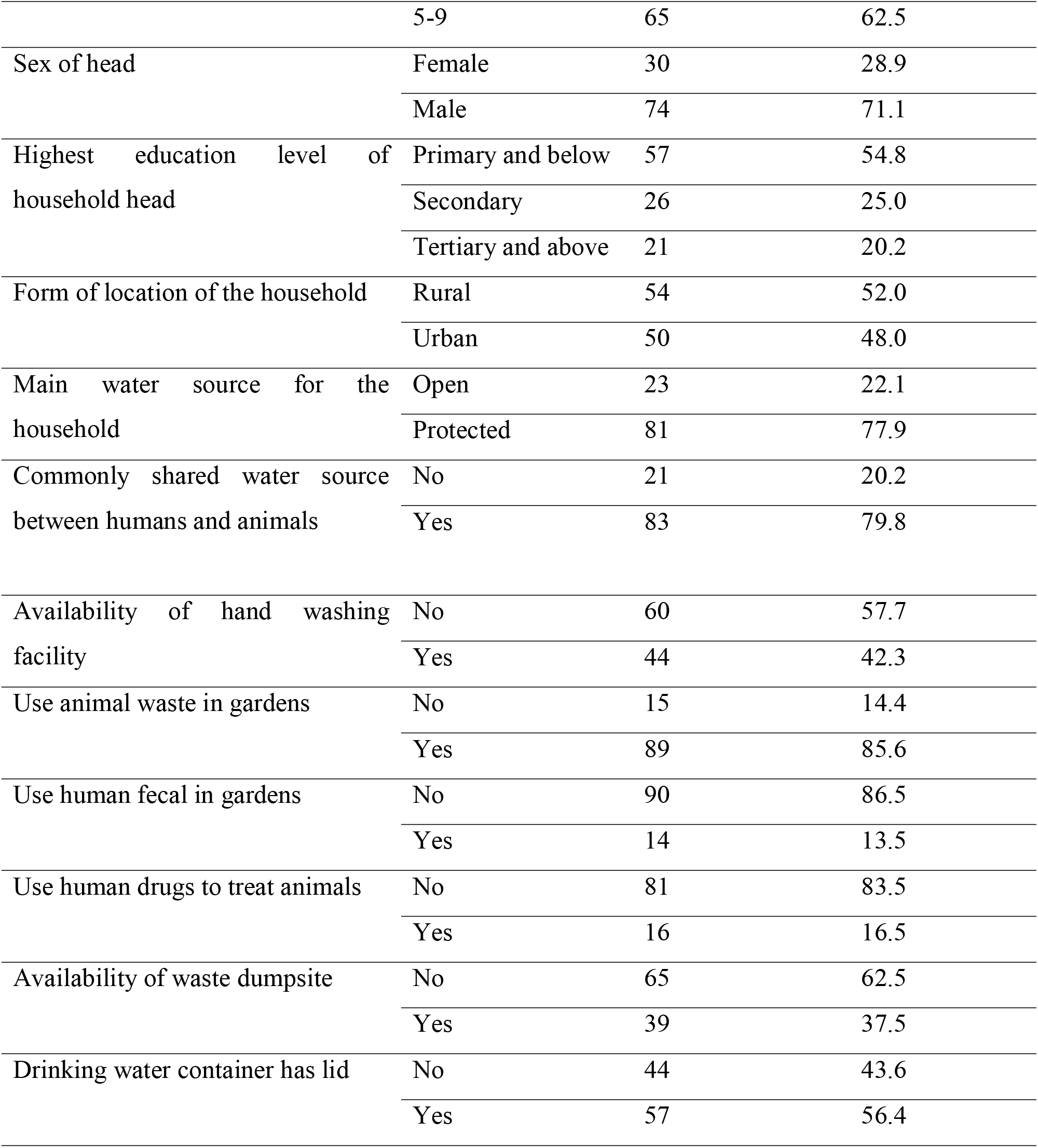
Characteristics of the study households.

### Characteristics of human and animal participants in the study

A total of 196 participants were recruited to provide samples that were used in this study. About 49.5% (97/196) of the study participants were aged below 18 years whereas just 10% (19/196) were aged between 18-35 years. Over 57.7% (113/196) of the participants were females. Additionally, 31.6% (62/196) of the participants were farmers. About 32.1% (63/196) and 20.9% (41/196) of the participants had been commonly suffering from intestinal and urinal related illnesses respectively. Samples were collected from a total of 393 animals in our study. Compared to other animals, samples were mostly collected from shoats 25.0% (99/393). Intensive animal husbandry was the predominant form of animal rearing 37% (145/393). Most 82% (321/393) of the animals were reported to be treated by a veterinarian. Details about the human and animal characteristics are given in **Error! Reference source not found**..

**Table Error!.** Reference source not found.: Characteristics of human and animal participants.

| Factors | Level | Frequency<br>(n=196) | Percentage<br>(%) |
| --- | --- | --- | --- |
| **Human participants** |  |  |  |
| Age | Less than 18 | 97 | 49.5 |
|  | 18-35 | 19 | 9.7 |
|  | Over 35 | 80 | 40.8 |
| Sex | Female | 113 | 57.7 |
|  | Male | 83 | 42.3 |
| Education level | Formal | 160 | 81.6 |
|  | None | 36 | 18.4 |
| Occupation | Farmer | 62 | 31.6 |
|  | None | 28 | 14.3 |
|  | Others | 26 | 13.3 |
|  | Student | 80 | 40.8 |
| Common condition you suffer from | Intestinal | 63 | 32.1 |
|  | Urinal | 41 | 20.9 |
|  | None | 92 | 46.9 |
| Sometimes did not complete treatment | No | 98 | 50.0 |
|  | Yes | 98 | 50.0 |
| Often closely interacted with animals | No | 32 | 16.5 |
|  | Yes | 162 | 83.5 |

| **Animal subjects** |  |  |  |
| --- | --- | --- | --- |
| Species | <sup>a</sup>Shoats | 99 | 25.2 |
|  | <sup>b</sup>Poultry | 92 | 23.4 |
|  | <sup>c</sup>Companion animals | 66 | 16.8 |
|  | Pig | 78 | 19.8 |
|  | Cattle | 58 | 14.8 |
| Husbandry practice | Backyard | 110 | 28.0 |
|  | Free range | 138 | 35.1 |
|  | Intensive | 145 | 36.9 |
| Feeding strategy | Kitchen leftover | 89 | 22.7 |
|  | Natural feeds (grass) | 254 | 64.6 |
|  | Supplementary feeds (mixed) | 50 | 12.7 |
| Source of water for animals | Protected | 276 | 70.2 |
|  | Unprotected | 117 | 29.8 |
| Treatment options | Experienced farmer | 5 | 1.3 |
|  | Owner treated | 67 | 17.1 |
|  | Veterinarian | 321 | 81.7 |

<sup>a</sup>Shoats meant sheep and goats; <sup>b</sup>Poultry meant turkey, chicken, and ducks; <sup>c</sup>Companion animals included: rabbits, cats and dogs

### Characteristics of environmental samples

A total of 399 environmental samples were collected from selected households. Approximately 25% (99/399), 25% (100/399), 26% (103/399) and 24% (97/399) samples were collected for water, soil, doorknobs, and animal feeders respectively. About 85% (85/99) of the water sources from which we sampled were shared between humans and animals. Nearly 65% (258/399) of the samples were obtained from soils that had human fecal matter and approximately 91% (364/399) had animals grazing on it.

### Burden (prevalence and severity) of ESBL-PE among households in Wakiso district

More than 80% (86/104) of the households had at least one positive sample. Up to 62% (95% CI: 51.5-70.8) of the sampled households were found to have humans carrying ESBL-PE. Over 70% (77/104) of the sampled households had animals carrying ESBL-PE. When assessed at sample level, the overall prevalence of ESBL-PE at the human-animal-environment interface among samples in Wakiso district was approximately 25% (95% CI: 22.7-28.3). At individual level, the animals, environment, and humans had ESBL-PE prevalence of 35.4%, 5.8%, and 45.4% respectively (**Error! Reference source not found**.3).

About 17.3% (18/104) of the sampled households had no positive ESBL-PE samples and were therefore was considered to be low risk. A similar number of households however presented with a high burden implying that all the three sampled components were found to be positive for ESBL-PE. Approximately 26.0% (27/104) of the households sampled had one of the components being positive (Figure).

**TableFigure :**
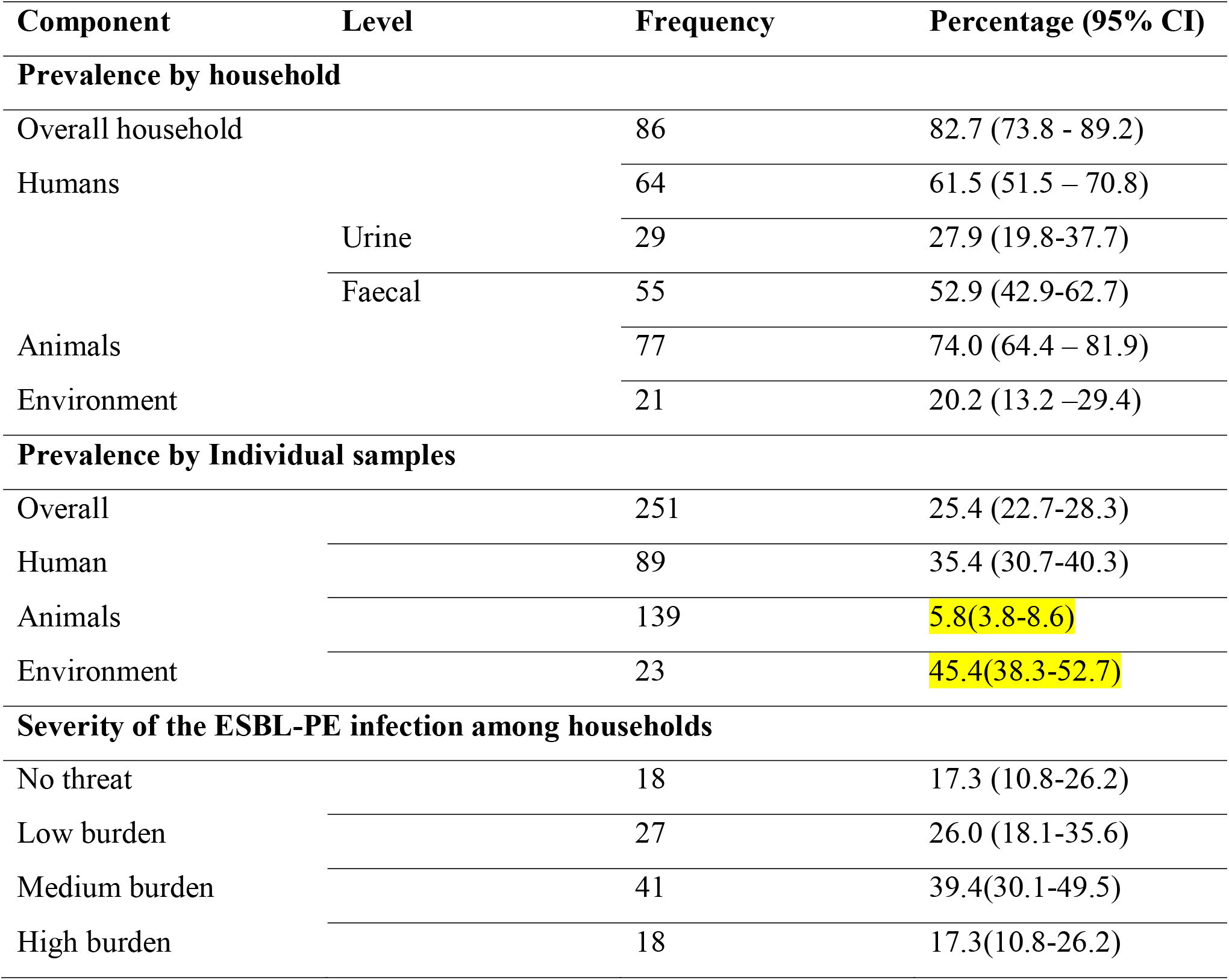
Burden of ESBL-PE among humans, animals, environment, and households.

### Distribution of ESBL-PE positivity among households

Figure 3 shows the different counts of households with the respective components. Majority of the positive households 45.3% (39/86) had both humans and animals were positive but not environment. Scenarios with only one component being positive yielded fewer representative households in the order (Animal>Human>Environment). About 17 households had all the 3 components being positive while no household had both the environment and humans being positive at the same time (Figure 3).

**Figure 3:**
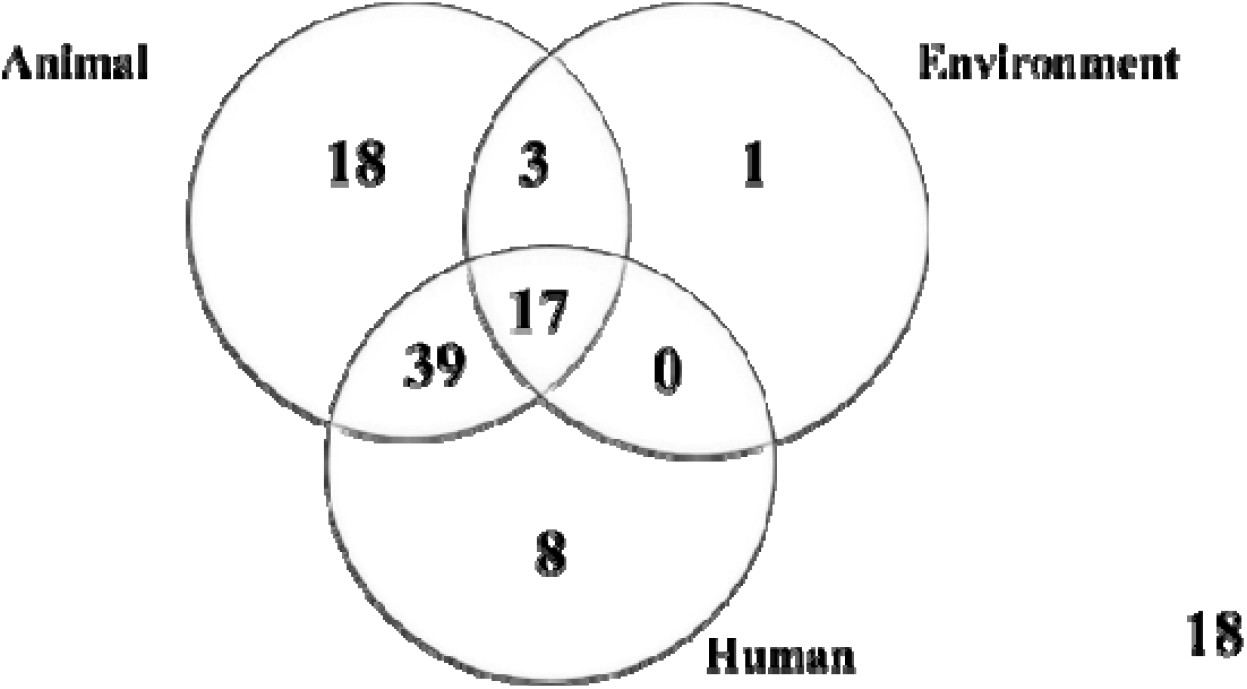
Distribution of ESBL-PE positivity among households.

### Factors associated with ESBL-PE human carriage and household contamination

Notably, households that had received a visitor in the last 24 hours had a higher ESBL PE prevalence among humans (adj PR=1.34, 95% CI: 1.13-1.60) compared to those that didn’t have a visitor. The prevalence of ESBL-PE was lower among those that collected water from protected water sources such as taps, springs compared to those that collected water from unprotected water sources such as wells (adj PR=0.63, 95%CI: 0.46-0.88). Participants in households where the drinking water was collected from a container with a lid had a lower prevalence of ESBL-PE (adj PR= 0.84 95% CI: 0.73-0.96), in addition, households whose environment was found dirty also had a lower burden of ESBL-PE among humans (adj PR=0.71 95%CI: 0.53-0.96) compared to the households with clean environments (Figure).

We found out that households that had a visitor in the last 24 hours were associated with high ESBL contamination (adj PR= 1.19, 95% CI: 1.04-1.36) compared to those that had no visitors. Notably, households that were using veterinary workers to administer drugs to their sick animal were significantly associated with a higher ESBL-PE contamination in households (adj PR= 1.39, 95% CI: 1.20-1.61) and, those that were using animal waste were more likely to be contaminated with ESBL-PE (adj PR= 1.29, 95% CI: 1.05-1.60). Additionally, households that reported getting medicines from professional health workers were more likely to be contaminated with ESBL-PE (adj PR=1.39, 95% CI: 1.20-1.61) compared to those that were getting from non-professionals (Figure 4).

**Table Figure :** Factors associated with ESBL-PE carriage and contamination among households.

| Variable | Level | Un adjusted PR (95% CI) | adjusted PR (95% CI) | p-value |
| --- | --- | --- | --- | --- |
| **Human ESBL-PE carriage** |  |  |  |  |
| Household that had a visitor in the last 24 hours | No | 1 | Ref |  |
|  | Yes | 0.92(0.81-1.03) | 1.34(1.13-1.60) | 0.0009*** |
| Protected Water source used as the main water source | No | 1 | Ref |  |
|  | Yes | **0.85(0.76-0.95) | 0.63(0.46-0.88) | 0.006** |
| Presence of a hand washing facility at home | No | 1 | Ref |  |
|  | Yes | *0.86(0.76-0.97) | 1.21(0.91-1.62) | 0.19 |
| Drugs administered to sick animals by a veterinary-personnel | No | 1 | Ref |  |
|  | Yes | **1.39(1.13-1.71) | 1.10(0.85-1.39) | 0.48 |
| Guidance on how long to wait after treating animal provided by veterinary personnel | No | 1 | Ref |  |
|  | Yes | 1.08(0.96-1.23) | 0.84(0.70-1.01) | 0.068 |
| Household compound found dirty | No | 1 | Ref |  |
|  | Yes | 0.93(0.83-1.05) | 0.71(0.53-0.96) | 0.024* |
| Presence of stagnant water | No | 1 | Ref |  |
|  | Yes | *0.88(0.76-1.00) | 0.93(0.70-1.23) | 0.59 |
| Drinking water container with lid | No | 1 | Ref |  |
|  | Yes | *0.87(0.78-0.98) | 0.84(0.73-0.96) | 0.014* |
| **Household ESBL-PE contamination** |  |  |  |  |
| Household had a visitor in the last 24 hours | No | 1 | 1 | Ref |
|  | Yes | 0.95(0.88-1.04) | 1.19(1.04-1.36) | 0.009** |
| Presence of household member who had been to another district or country in the past one month | No | 1 | 1 | Ref |
|  | Yes | *1.08(1.01-1.16) | 1.11(0.94-1.30) | 0.23 |
| Source of drugs | Nonprofessional | 1 | 1 | Ref |
|  | Professional | *1.15(0.99-1.33) | 1.39(1.20-1.61) | <0.05*** |
| Reasons for negotiating on treatment | Financial | 1 | 1 | Ref |
|  | Side effects | 1.09(0.97-1.23) | 1.09(1.00-1.19) | 0.06 |
| Cleaning of animal house | Twice a week | 1 | 1 | Ref |
|  | Once a week | *1.10(1.00-1.19) | 1.06(0.95-1.18) | 0.25 |
|  | Throughout a week | 0.97(0.87-1.07) | 0.95(0.84-1.07) | 0.41 |
| Management of animal waste | Dispose | 1 | 1 | Ref |
|  | Use | 1.11(0.96-1.29) | 1.29(1.05-1.60) | 0.02* |
| Drugs administered to sick animals by veterinary personnel | No | 1 | 1 | Ref |
|  | Yes | *1.28(1.05-1.58) | 1.39(1.20-1.61) | <0.05** |

## DISCUSSION

The burden of ESBL-PE carriage among apparently healthy individuals and animals has been reported globally with little evidence on the environment [8]. There is scanty information about the ESBL-PE among humans, animals and their immediate environment especially in the LMICs such as Uganda. Surveillance of ESBL-PE “globally recognized sentinel organism for AMR” at the human-animal-environment interface provides information required to design sustainable one health interventions globally. In our study, we report a high household ESBL-PE burden. Having an animal health worker treat household animals and obtaining medical attention from a health facility were associated with a high prevalence of ESBL-PE carriage and contamination among humans and household respectively. These two results are indicative of the poor infection prevention and control measures by both the human and animal health workers thus presenting a great concern for public health.

This study reports an overall ESBL-PE prevalence of 25.4% from all the components sampled (i.e. humans, animals and environment). On the contrary, authors in Nepal reported 53.4% prevalence of ESBL-PE at the human-animal-environment interface [9]. This difference could potentially be attributed to the variation in environmental sampling in the two studies. The earlier study considered community drainage and sewage systems round households which overestimated the ESBL-PE prevalence at household level unlike the current one that utilized the immediate household environment that yielded a comparatively low ESBL-PE. Over 83% of the sampled households had a positive ESBL-PE sample irrespective of its source. Such findings reveal a high level of household contamination and risk of ESBL-PE infections especially in a typical farming community. In case of an infection and disease, this situation will certainly drive the cost of treatment high, long stay in the hospitals as well as compromised quality of health outcomes [10].

Majority of the households had animals contributing highest to ESBL-PE household positivity followed by humans and environment being least contributor. This result could be attributed to the fact that *E. coli* is a harmless inhabitant of the human and animal guts [11] apart from its key role in the transmission of ESBL genes. The regular and irrational use of antimicrobials in animal production could also trigger the development of ESBL-PE in animals [12]. Several human related factors such as self-medication, buying drugs over the counter, animal food products containing drug residues and the general transmission dynamics of the organisms could explain the observed burden among humans [13]. Perhaps the low contribution to environment ESBL-PE prevalence is because the environment subjects the bacteria to harsh conditions and therefore their population is constantly being checked [29,30]. Studies that have documented a high burden of ESBL-PE in the environment have sampled the sites with constant supply of nutrients for the survival of the organisms such as storm water drains and sewerage systems [31,32]. Our study utilized the immediate environment of humans and animals to get a clear indicator of transmission dynamics of ESBL-PE at household level. It is therefore important that future studies focus on the survival and viability of ESBL-PE in environmental components exposed to harsh weather conditions such as sunshine and heavy rainfall.

Interestingly, humans from households that had a visitor in the last 24 hours registered a high ESBL-PE prevalence. Similarly, household that had a visitor in the last 24 hours had a higher ESBL-PE contamination compared to those that never had a visitor. Indeed, visiting has been earlier implicated in not only ESBL-PE occurrence but also the general AMR transmission cycle [33,34]. This could potentially be attributed to the fact that visitors come from different locations, use different transport means and might be carrying resistant organisms such as ESBL-PE. Infection prevention and control strategies such as hand washing with soap and sanitizing are crucial for households especially when welcoming visitors.

Important to note, obtaining water from a protected source was associated with a low ESBL-PE carriage among humans. Protected water sources provide wholesome water free form contamination by humans, animals, and storm water [20]. Unprotected water sources are vulnerable to a wide range of contamination including ESBL-PE from the environment and anthropogenic activities [21]. In line with the safe water chain, our study revealed that having drinking water containers covered with lids was associated to low ESBL-PE human carriage. Safe water storage reduces the bacterial load and further contamination is prevented by covering the storage container with a lid [22]. A similar study done Brick and colleagues in 2004 revealed that covering drinking water container with a lid limited contamination [23]. Therefore, there is need for communities to ensure a safe water chain from the water source up to use.

Interestingly, humans from households with dirty compounds were found to have less ESBL-PE carriage. A study conducted on improving surface sampling and detection of contamination presents different views and contentions around dirty environments and safety [24]. This could be due to a number of factors such as use of personal protective equipment (PPE), low survival of ESBL-PE in the harsh environment, proper personal hygiene practices, among others in farming communities. Several studies are in agreement with good hygienic practices, increased household use of PPE and low abundance of bacterial pathogens in the environment could potentially lead to reduced contamination especially with ESBL-PE organisms [40,41]. Therefore, more environmental studies are required on bacterial abundance and AMR especially in household environments.

The households that got the drugs from the professional healthcare providers were more likely to be contaminated with ESBL-PE. Given the high burden of drug resistant organisms, household members who attend or seek medical attention from health care facilities are at a higher risk of contracting nosocomial infections most of which are bacteria in nature such as ESBL-PE [27]. Humans have ability to shed the contracted bacteria into the environment and or with the animals with in the household setting [13]. Therefore, it is important to improve the infection prevention and control in the health facilities in order to minimize hospital acquired infections among community members.

Households that contacted a veterinary worker to administer the drugs to their sick animal were more likely to be at risk of ESBL-PE contamination. Poor on farm biosafety measures, improper compliance to the use of PPE, reuse of veterinary equipment among farms, poor storage of veterinary drugs among others could potentially contribute to the reported household contamination by veterinary workers [28]. In addition, misuse of veterinary products without prior diagnosis could escalate resistance among animals in households [29]. Therefore, improved on farm biosafety measures, good veterinary practices as well as veterinary drug use regulations are paramount to reverse the problem of AMR.

Households that use the animal waste in several activities such as farming, biogas production were more likely to be contaminated with ESBL-PE than those that dispose of the waste. Studies have indicated the presence of ESBL-PE in animal waste [26]. Utilization of such waste without decreasing the contamination load and without biosafety precautions exposes the human, other animals and the environment to subsequent ESBL-PE contamination [30]. Therefore, the use of PPE during waste handling, decontamination of animal waste before use should be emphasized among farming households in order to curb contamination and ESBL-PE transmission.

## Conclusion

This study is among the first of its kind in Uganda focusing at determining the ESBL-PE carriage, contamination, and associated risk factors at household level among farming communities. A wide spread of ESBL-PE among humans, environment and animals indicates a great public health threat. This observation could potentially be due to poor infection prevention and control (IPC) measures in the area by veterinarians and household members. The role of human, environment, and animal health workers in the occurrence of ESBL-PE and other types of AMR is therefore critical. Improved collaborative AMR mitigation strategies such as safe water chain, on farm biosecurity, household, and facility-based IPC measures as well as capacity building of the human, animal and environmental health workers using a one health approach is paramount in order to curb the problem of AMR among farming communities.

## Data Availability

All data produced in the present study are available upon reasonable request to the authors

## List of abbreviations

AMR: Antimicrobial resistance
ARP: Antimicrobial Resistant pathogens
*E. coli*: *Escherichia coli*
ESBL-PE: Extended Spectrum Beta Lactamase Producing-*E. coli*
IPC: Infection Prevention and Control
LMICs: Low-and middle-income countries
NAP: National Action Plan for Antimicrobial Resistance
PPE: Personal Protective Equipment
WHO: World Health Organization

## Funding statement

*“This research (or “[initials of fellow]”) was supported by the Consortium for Advanced Research Training in Africa (CARTA). CARTA is jointly led by the African Population and Health Research Center and the University of the Witwatersrand and funded by the Carnegie Corporation of New York (Grant No. G-19-57145), Sida (Grant No:****54100113****), Uppsala Monitoring Center, Norwegian Agency for Development Cooperation (Norad), and by the Wellcome Trust [reference no. 107768/Z/15/Z] and the UK Foreign, Commonwealth & Development Office, with support from the Developing Excellence in Leadership, Training and Science in Africa (DELTAS Africa) programme. The statements made and views expressed are solely the responsibility of the Fellow. In addition, CIDIMOH project under the NORHED II funding supported part of the laboratory component during analysis*.

### Availability of data and materials Data will be availed upon request Author contributions

JM, SK, FEM, EB, JCS, CK, MM and DM BEB conceptualised and designed the study. JM, DM, JCS, CK, JBI, RW supervised and coordinated the data collection. JM and RKM, RW supported and coordinated data collection. JM, FEM, SK, BEB carried out data analyses. JM, RW, JCS, DM, CK, MM, JBI, RKM drafted the initial manuscript, and all authors approved the final manuscript as submitted. All authors read and approved the final manuscript.

## Corresponding author

Correspondence to James Muleme

## Acknowledgments

We are grateful to the respondents who participated in this study. We also thank the district and sub county officials who supported the study inception and processes. The local authorities in the respective study sites were very supportive and our research participants in the same regard.

